# World Science against COVID-19: Gender and Geographical Distribution of Research

**DOI:** 10.1101/2021.09.29.21264261

**Authors:** Julio González-Álvarez

## Abstract

In just a year and a half, an enormous volume of scientific research has been generated throughout the world to study a virus/disease that turned into a pandemic. All the articles on COVID-19 or SARS-CoV-2 included in the SCI-EXPANDED database (Web of Science), signed by more than a third of a million of authorships, were analyzed. Gender could be identified in 92% of the authorships. Women represent 40% of all authors, a similar proportion as first authors, but just 30% as last/senior authors. The pattern of collaboration shows an interesting finding: when a woman signs as a first or last/senior author, the article byline approximates gender parity

According to the corresponding address, the USA shares 22.8% of all world articles, followed by China (14.4%), Italy (7.8%), the UK (5.8%), India (4.2%), Spain (3.8%), Germany (3.6%), France (2.9%), Turkey (2.5%), and Canada (2.4%).

Despite their short lives, the papers received an average of 11 citations. The high impact of papers from China is striking (25.1 citations; the UK, 12.4 citations; the USA, 11.3 citations), presumably because the disease emerged in China, and the first publications (very cited) came from there.

---

In December 2019, the Chinese city of Wuhan became the center of an outbreak of pneumonia of unknown origin. One month later, Chinese scientists isolated a novel coronavirus, the severe acute respiratory syndrome coronavirus 2, or SARS-CoV-2, responsible for this viral pneumonia, which was later designated coronavirus disease 2019 (COVID-19) by the World Health Organization^1^.

Since then, in just a year and a half, an enormous volume of scientific research has been generated throughout the world to study this new virus/disease turned into a pandemic. I analyzed all the articles on COVID-19 or SARS-CoV-2 included in the Science Citation Index Expanded (SCI-EXPANDED) database of Web of Science, signed by more than a third of a million of authorships. The analysis was done from a double perspective as I wanted to determine the gender composition of the authors and discover the participation of women in this gigantic scientific endeavor. Furthermore, I wanted to know the participation of the different regions of the world by analyzing the corresponding addresses.

## Method

### Sample data

All the Articles on COVID-19 or SARS-CoV-2 (TOPIC) included in the SCI-EXPANDED database (Web of Science, Clarivate Analytics) were selected on 10–13 May 2021. It is recognized that this database includes the world’s leading journals of science and technology after a rigorous selection process Our collection consisted of 40,765 articles signed by 340,868 authorships and published in 3,810 journals. The articles were published in 2021 (16,821), 2020 (23,941), and 2019 (3). See methodological details in the supplementary material.

### Gender identification of authors

I examined the authorships to determine their gender. The SCI-EXPANDED database (like most scientific database) does not provide information about the authors’ gender. However, in 2008 the Web of Science began to include the authors’ full names, although a small proportion of records still display only the authors’ initials. All the authors’ first names were matched through two gender databases: GenderChecker (acquired from http://genderchecker.com/) and Gender API (acquired from https://genderapi.io/).

### Procedure

Each variable of interest (author name and surnames, title of article, year of publication, journal, corresponding address, etc.) was extracted using the BibExcel program^2^ and merged in a master Excel database to perform the bibliometric analyses. Statistical analyses were carried out with the SPSS v.22 software.

## Results and Discussion

### Rate of women authors

From the total 340,868 authorships, and after excluding the authorships with only initials, unisex names, or first names that did not match the gender databases, gender could be identified in 314,319 (92.2%). Men were 188,465, and women, 125,854. Therefore, women represent **40%** of all the known-gender authorships^a^. This percentage of female researchers regarding COVID-19 (or SARS-CoV-2) is quite far from the gender parity [X^2^(df = 1) = 6297.99, p< .0001, Cramer’s V = 0.10^b^], although a somewhat larger proportion than the overall presence of women in worldwide science, about a third of researchers^3^. González-Alvarez analyzed *The Lancet* journals during 2014–17 and found that women only represented about one-third of the authors in *The Lancet* (31.8%) and six other *Lancet* journals, whereas *The Lancet Psychiatry* (45.2%), *The Lancet Global Health* (39.8%), and *The Lancet HIV* (38.8%) presented a somewhat lower gender imbalance^4^.

I identified the order of signature in all articles on COVID-19 and obtained the gender percentages as first and last authors. The first and last (or senior) places are usually key positions in health and medical sciences. In relative terms, women are slightly underrepresented as first authors, totaling **38.6**% of them (Table 1, last row; Figure 1). Filardo et al.^5^ observed that female first authorship increased significantly from 27% in 1994 to 37% in 2014 for articles published in six high-impact medical journals; that proportion of 37% is close to ours 38.6% for all articles on COVID-19. However, Gonzalez-Alvarez and Sos-Peña^6^ very recently studied 40 journals of different impact factors published in Cancer research and found that the proportion of women as first authors increased to 43.8%.

**Table 1.**
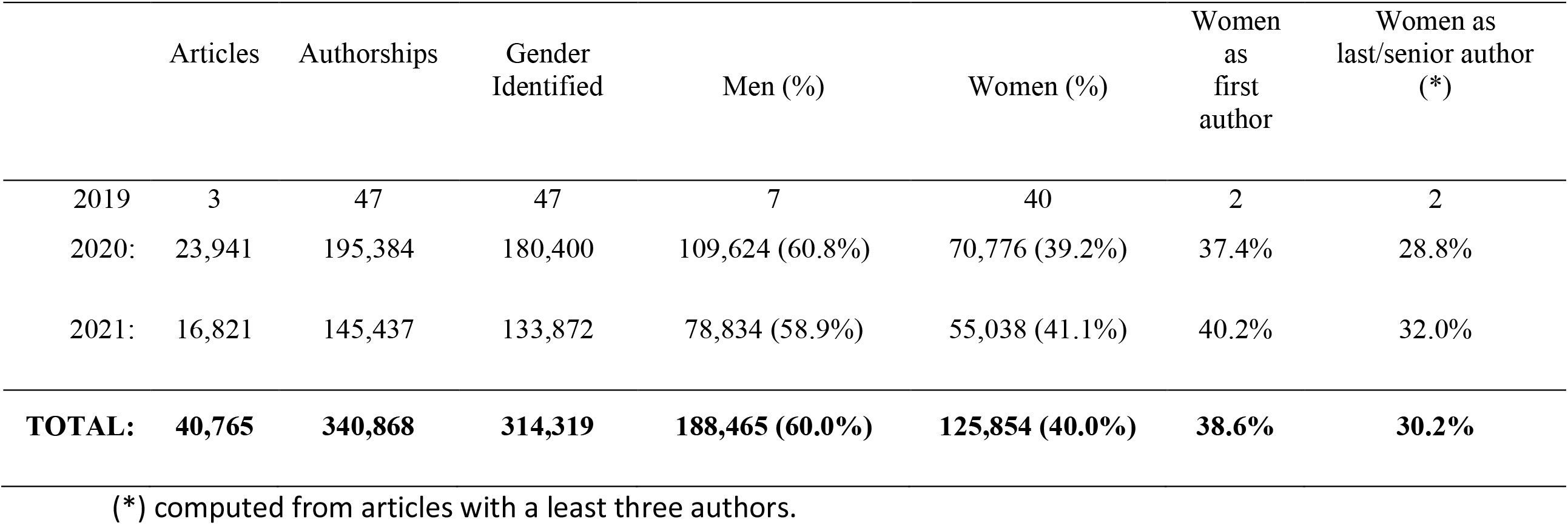
Data from all the Articles on **COVID-19** or **SARS-CoV-2** obtained from the **SCI-EXPANDED** (Science Citation Index Expanded) from the Web of Science (Clarivate Analytics).

|  | Articles | Authorships | Gender Identified | Men (%) | Women (%) | Women as first author | Women as last/senior author (*) |
| --- | --- | --- | --- | --- | --- | --- | --- |
| 2019 | 3 | 47 | 47 | 7 | 40 | 2 | 2 |
| 2020: | 23,941 | 195,384 | 180,400 | 109,624 (60.8%) | 70,776 (39.2%) | 37.4% | 28.8% |
| 2021: | 16,821 | 145,437 | 133,872 | 78,834 (58.9%) | 55,038 (41.1%) | 40.2% | 32.0% |
| **TOTAL:** | **40,765** | **340,868** | **314,319** | **188,465 (60.0%)** | **125,854 (40.0%)** | **38.6%** | **30.2%** |

(\*) computed from articles with a least three authors.

**Figure 1.**
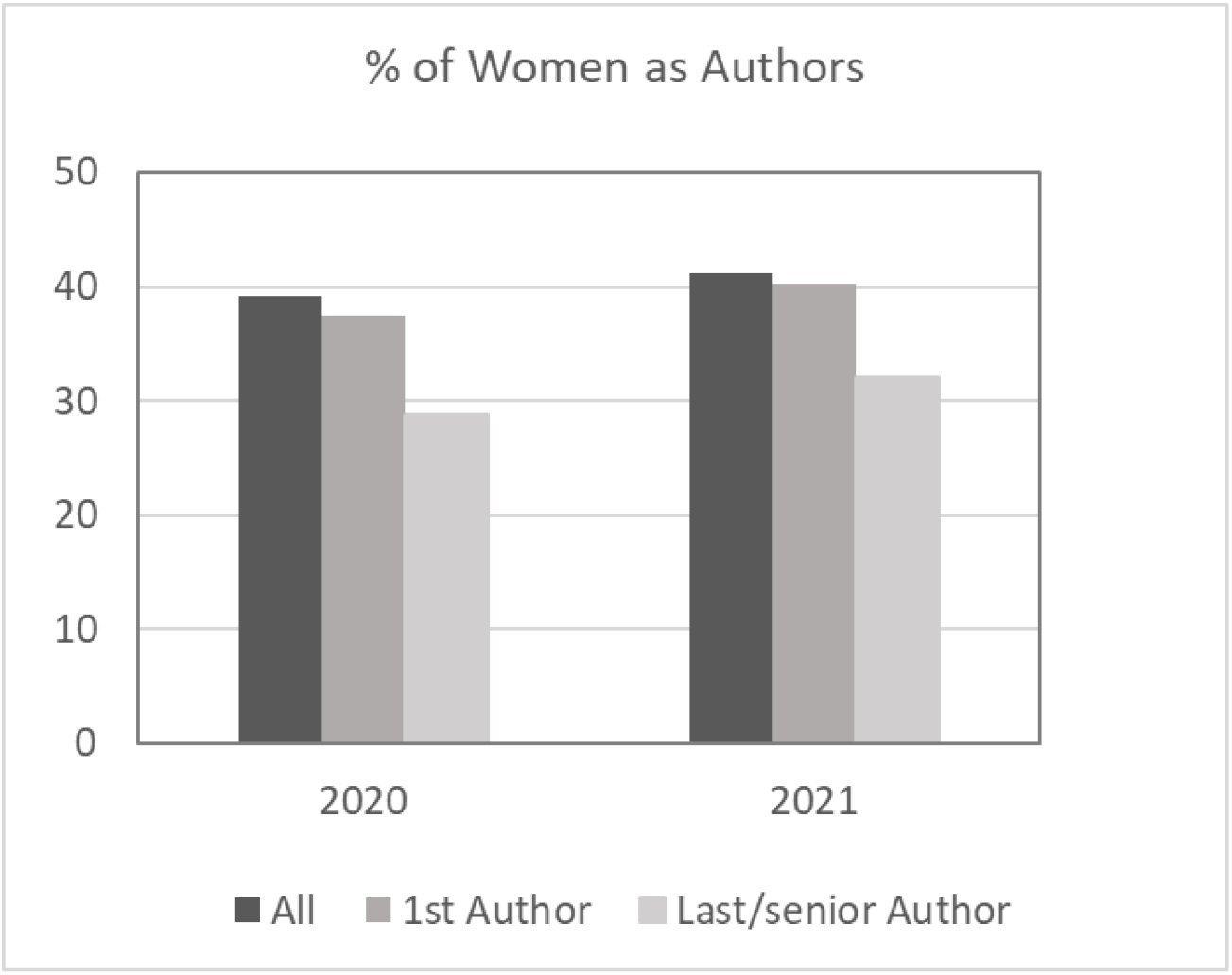
Percentages of women as authors of all the articles published in 2020/21 on COVID-19 or SARS-CoV-2 (SCI-EXPANDED, Web of Science). The figure displays overall percentages and percentages of women as first or last/senior authors.

Our data show that women are clearly underrepresented as last/senior authors compared to the overall rate: only **30.2**% of the last authors of articles on COVID-19, signed by three or more authors, were women (Table 1, Figure 1). In biomedical sciences, this position is usually reserved for the senior or leading scientist on a research project and normally corresponds to a scientist with a consolidated and longer career^7^. This relative female underrepresentation as last/senior authors has also been observed in other gender studies on scientific and biomedical publications^3,4,6^, suggesting that, in addition to other variables, age—or more exactly, seniority—might play some role in the gender composition of COVID-19 researchers. This may point toward an optimistic scenario in the sense that gender imbalance could be reduced as new generations of female researchers are gaining seniority.

### The pattern of collaboration

Research for COVID-19 seems a very collaborative field, with great differences in the number of authors per paper. The articles were written by an average of 8.4 authors, with a range from an article published in *The Lancet* signed by 2972 authors (belonging to the RECOVERY Collaborative Group), or 26 articles with more than 100 authors, to almost 2000 articles signed by a single author.

Taking into account the gender of the authors, I considered the articles with at least three co-authors divided into two groups (as in González-Álvarez & Sos-Peña^6^): a) articles signed by a man as the first author and b) articles signed by a woman as the first author. I repeated the procedure and divided the initial set of articles into two new groups: c) articles signed by a man as the last/senior author and d) articles signed by a woman as the last/senior author. Analyzing the gender composition of each group, I observed a significant finding (Figure 2), also found in contemporary research on cancer^6^. **The articles signed by a woman in the first or last/senior position approach gender parity in the byline** (48.1% men/51.9% women when a woman signed as the first author, and 45.8% men/54.2% women when a woman signed as the last/senior author, which was slightly female-biased). This fact does not necessarily mean that there is a cause-and-effect relationship between the presence of women in one of these two key positions and near gender parity in the article byline, but these two facts are correlated. It gives the impression that leading female researchers tend to co-publish with women more than leading male researchers do; alternatively, they may be working on subtopics that are relatively more appealing to women.

**Figure 2.**
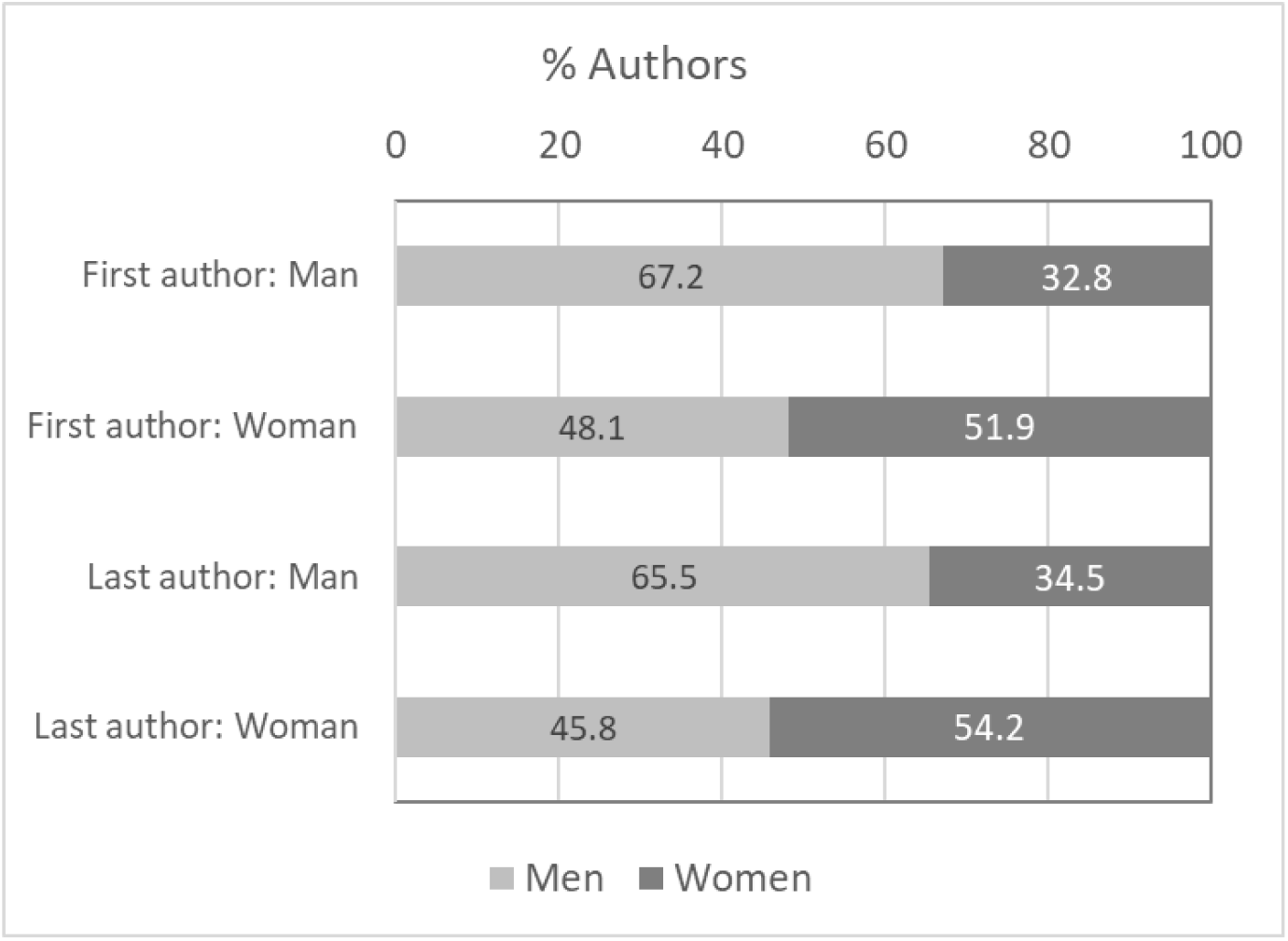
Percentages of Men and Women as authors, depending on which gender occupied the first or last/senior positions in the article byline. Values were calculated for articles with at least three coauthors.

On the contrary, when the first author is a man, the gender composition of the byline is more asymmetrical (67.2% men/32.8% women, Figure 2), compared to the overall asymmetry (60.0% men/40.0% women), *χ*^2^(df=1) = 2576.77, p < .0001, Cramer’s V = 0.07. The same pattern appears when a man is the last or senior author (65.5% men/34.5% women, Figure 2), *χ*^2^(df=1) = 1655.57, p < .0001, Cramer’s V = 0.06.

### Geographical distribution

Although research on COVID-19 is logically transnational in most groups, I considered the corresponding address of each article. Table 2 shows data from the 20 countries with the highest number of articles. In Table S1 of the supplementary material, we can see the list complete of all the countries. According to the corresponding addresses, the United States of America (USA) shares 22.8% of all world articles, followed by China (14.4%), Italy (7.8%), the United Kingdom (UK) (5.8%), India (4.2%), Spain (3.8%), Germany (3.6%), France (2.9%), Turkey (2.5%), and Canada (2.4%). Among the 10 most productive countries, India stands out for its greater gender inequality (only 29.2% of the researchers are women), and, at the other extreme, Spain stands out for its larger gender balance (46.5% female researchers).

**Table 2.** Data from the twenty countries (according to the corresponding address) with the highest number of articles.

|  |  | Articles | % | Authorships | Authors/<br>Article | Known<br>Gender | Men | % | Women | % | Citations<br>received | Citations/<br>Article |
| --- | --- | --- | --- | --- | --- | --- | --- | --- | --- | --- | --- | --- |
| 1 | USA | **9274** | **22.8** | 75875 | 8.18 | 73267 | 42665 | 58.2 | 30602 | 41.8 | 104720 | 11.29 |
| 2 | Peoples R China | **5861** | **14.4** | 54342 | 9.27 | 47654 | 28229 | 59.2 | 19425 | 40.8 | 147168 | 25.11 |
| 3 | Italy | **3174** | **7.8** | 33297 | 10.49 | 31478 | 17978 | 57.1 | 13500 | 42.9 | 32241 | 10.16 |
| 4 | UK | **2372** | **5.8** | 24732 | 10.43 | 20298 | 11930 | 58.8 | 8368 | 41.2 | 29461 | 12.42 |
| 5 | India | **1703** | **4.2** | 10104 | 5.93 | 8864 | 6277 | 70.8 | 2587 | 29.2 | 8534 | 5.01 |
| 6 | Spain | **1529** | **3.8** | 14091 | 9.22 | 13178 | 7051 | 53.5 | 6127 | 46.5 | 9223 | 6.03 |
| 7 | Germany | **1458** | **3.6** | 13818 | 9.48 | 12680 | 8675 | 68.4 | 4005 | 31.6 | 11615 | 7.97 |
| 8 | France | **1189** | **2.9** | 13603 | 11.44 | 12192 | 7556 | 62.0 | 4636 | 38.0 | 15215 | 12.80 |
| 9 | Turkey | **1011** | **2.5** | 5927 | 5.86 | 5744 | 3476 | 60.5 | 2268 | 39.5 | 3251 | 3.22 |
| 10 | Canada | **972** | **2.4** | 7002 | 7.20 | 6688 | 3848 | 57.5 | 2840 | 42.5 | 7054 | 7.26 |
| 11 | Brazil | **920** | **2.3** | 7168 | 7.79 | 6928 | 3704 | 53.5 | 3224 | 46.5 | 4382 | 4.76 |
| 12 | Australia | **836** | **2.1** | 6196 | 7.41 | 5946 | 3440 | 57.9 | 2506 | 42.1 | 6021 | 7.20 |
| 13 | South Korea | **659** | **1.6** | 4448 | 6.75 | 4208 | 2737 | 65.0 | 1471 | 35.0 | 5334 | 8.09 |
| 14 | Japan | **648** | **1.6** | 5217 | 8.05 | 5046 | 3935 | 78.0 | 1111 | 22.0 | 3946 | 6.09 |
| 15 | Iran | **620** | **1.5** | 4249 | 6.85 | 4099 | 2558 | 62.4 | 1541 | 37.6 | 3349 | 5.40 |
| 16 | Saudi Arabia | **462** | **1.1** | 2794 | 6.05 | 2626 | 1926 | 73.3 | 700 | 26.7 | 1782 | 3.86 |
| 17 | Switzerland | **399** | **1.0** | 3230 | 8.10 | 3058 | 1906 | 62.3 | 1152 | 37.7 | 3973 | 9.96 |
| 18 | Netherlands | **363** | **0.9** | 3057 | 8.42 | 2740 | 1615 | 58.9 | 1125 | 41.1 | 5549 | 15.29 |
| 19 | Israel | **348** | **0.9** | 2318 | 6.66 | 2249 | 1367 | 60.8 | 882 | 39.2 | 1687 | 4.85 |
| 20 | Singapore | **330** | **0.8** | 2933 | 8.89 | 2802 | 1729 | 61.7 | 1073 | 38.3 | 6674 | 20.22 |

### Citations

The number of citations received by each article was obtained. Despite their short lives, the papers received an average of 11 citations (Table 2, see the last two columns).The high impact of papers from China is striking (25.1 citations, compared to the UK: 12.4 citations, or the USA: 11.3 citations), presumably because the disease emerged in China, and the first publications (very cited) came from there.

Regarding gender, I assigned the citations of each article to each of its authors and compared the citations received by each gender. Men received a mean of 18,6 citations^c^, (SD = 106,5), 95% CI [18.1, 19.1]. Women received a mean of 17,0 citations, (SD = 100,6), 95% CI [16.5, 17.6]. The difference between the citations was significant, given the huge number of observations, F (1) = 16.65, MSe = 10854.66, p <.0001, but the effect size (η^2^_p_ = .000052^d^) was minimal.

Table 3 shows the 30 most cited articles on COVID-19. The list is headed by the paper “Clinical Characteristics of Coronavirus Disease 2019 in China” published in *The New England Journal of Medicine* by researchers from the *China Medical Treatment Expert Group for Covid-19*, and in just over a year, has received 8313 citations. This article is followed by “Clinical course and risk factors for mortality of adult inpatients with COVID-19 in Wuhan, China: a retrospective cohort study” published in *The Lancet* by Chinese researchers and receiving 7396 citations. At a considerable distance (2731 citations) this is followed by “Pathological findings of COVID-19 associated with acute respiratory distress syndrome” published in *The Lancet Respiratory Medicine* by researchers from different institutions of Beijing.

**Table 3.** The 30 most cited articles on COVID-19 or SARS-CoV-2 (SCI-EXPANDED, Web of Science).

| rank | Title | Year | Journal | Corresponding address | Citations received |
| --- | --- | --- | --- | --- | --- |
| 1 | Clinical Characteristics of Coronavirus Disease 2019 in China | 2020 | NEW ENGLAND JOURNAL OF MEDICINE | Peoples R China | **8313** |
| 2 | Clinical course and risk factors for mortality of adult inpatients with COVID-19 in Wuhan, China: a retrospective cohort study | 2020 | LANCET | Peoples R China | **7396** |
| 3 | Pathological findings of COVID-19 associated with acute respiratory distress syndrome | 2020 | LANCET RESPIRATORY MEDICINE | Peoples R China | **2731** |
| 4 | Presenting Characteristics, Comorbidities, and Outcomes Among 5700 Patients Hospitalized With COVID-19 in the New York City Area | 2020 | JAMA-JOURNAL OF THE AMERICAN MEDICAL ASSOCIATION | USA | **2084** |
| 5 | Risk Factors Associated with Acute Respiratory Distress Syndrome and Death in Patients with Coronavirus Disease 2019 Pneumonia in Wuhan, China | 2020 | JAMA INTERNAL MEDICINE | Peoples R China | **2052** |
| 6 | Hydroxychloroquine and azithromycin as a treatment of COVID-19: results of an open-label non-randomized clinical trial | 2020 | INTERNATIONAL JOURNAL OF ANTIMICROBIAL AGENTS | France | **2046** |
| 7 | Antibody responses to SARS-CoV-2 in patients with COVID-19 | 2020 | NATURE MEDICINE | Peoples R China | **2021** |
| 8 | A Trial of Lopinavir-Ritonavir in Adults Hospitalized with Severe Covid-19 | 2020 | NEW ENGLAND JOURNAL OF MEDICINE | Peoples R China | **1882** |
| 9 | Neurologic Manifestations of Hospitalized Patients with Coronavirus Disease 2019 in Wuhan, China | 2020 | JAMA NEUROLOGY | Peoples R China | **1761** |
| 10 | The species Severe acute respiratory syndrome-related coronavirus: classifying 2019-nCoV and naming it SARS-CoV-2 | 2020 | NATURE MICROBIOLOGY | Russia | **1747** |
| 11 | Immediate Psychological Responses and Associated Factors during the Initial Stage of the 2019 Coronavirus Disease (COVID-19) Epidemic among the General Population in China | 2020 | INTERNATIONAL JOURNAL OF ENVIRONMENTAL RESEARCH AND PUBLIC HEALTH | Singapore | **1614** |
| 12 | Correlation of Chest CT and RT-PCR Testing for Coronavirus Disease 2019 (COVID-19) in China: A Report of 1014 Cases | 2020 | RADIOLOGY | Peoples R China | **1555** |
| 13 | Baseline Characteristics and Outcomes of 1591 Patients Infected With SARS-CoV-2 Admitted to ICUs of the Lombardy Region, Italy | 2020 | JAMA-JOURNAL OF THE AMERICAN MEDICAL ASSOCIATION | Italy | **1542** |
| 14 | Incidence of thrombotic complications in critically ill ICU patients with COVID-19 | 2020 | THROMBOSIS RESEARCH | Netherlands | **1373** |
| 15 | Severe acute respiratory syndrome coronavirus 2 (SARS-CoV-2) and coronavirus disease-2019 (COVID-19): The epidemic and the challenges | 2020 | INTERNATIONAL JOURNAL OF ANTIMICROBIAL AGENTS | Taiwan | **1337** |
| 16 | The Incubation Period of Coronavirus Disease 2019 (COVID-19) From Publicly Reported Confirmed Cases: Estimation and Application | 2020 | ANNALS OF INTERNAL MEDICINE | USA | **1321** |
| 17 | Factors Associated with Mental Health Outcomes Among Health Care Workers Exposed to Coronavirus Disease 2019 | 2020 | JAMA NETWORK OPEN | Peoples R China | **1296** |
| 18 | Clinical characteristics and intrauterine vertical transmission potential of COVID-19 infection in nine pregnant women: a retrospective review of medical records | 2020 | LANCET | Peoples R China | **1249** |
| 19 | Structural basis for the recognition of SARS-CoV-2 by full-length human ACE2 | 2020 | SCIENCE | Peoples R China | **1242** |
| 20 | Clinical characteristics of 113 deceased patients with coronavirus disease 2019: retrospective study | 2020 | BMJ-BRITISH MEDICAL JOURNAL | Peoples R China | **1236** |

## Conclusions

After analyzing all the scientific articles on COVID-19/SARS-CoV-2 included in the SCI-EXPANDED database, we can draw the following conclusions:

- Women represent **40%** of authorships, still far from gender parity.
- Compared to the overall rate, women are relatively underrepresented as **last or senior** authors (30.2%). This fact, also found in other studies, suggests that age, or more specifically, seniority, could play some role in the gender composition of biomedical researchers.
- The pattern of collaboration shows an interesting finding, also observed in another study^6^: when a woman signs as the first or last/senior author, the article byline approximates gender parity.
- Considering the corresponding addresses, the largest volume of research on COVID-19 corresponds to the United States of America (22.8% of all articles), followed by China (14.4%).
- Despite their short lives (a year and a half), the articles have received an average of 11 citations. The high impact of papers from China is remarkable (25.1 citations), presumably because the disease emerged in China, and the first publications (very cited) came from there.

## Data Availability

Supplementary Information and Data available at the following link:

http://www.langproc.uji.es/COVID/World_Science_against_COVID.htm

## Ethical approval

This work did not require ethical approval.

## Acknowledgements

This work was completed with resources provided by the University Jaume I of Castellon (Spain).

## Footnotes

a The percentages of female or male authorships will always refer to the known-gender totals.

b Cramer’s V determines the effect size. The standard interpretation for one degree of freedom (df) is: 0.10 = small, 0.30 = medium, 0.50 = large effect.

c Each gender received more citations than the overall mean because the most cited articles tended to be signed by more authors.

d The effect size interpretations for partial eta squared (η^2^p) values are: .01 = small, .06 = medium, and .14 = large.

