## Supplementary Information for "World Science against COVID-19: Gender and Geographical Distribution of Research"

### Method

Records extracted on 10-13 May 2021 from the **SCI-EXPANDED** (Science Citation Index Expanded) from the Web of Science (Clarivate Analytics)

All the Articles on **COVID-19** or **SARS-CoV-2** were obtained.

SEARCH:

TOPIC: (COVID-19) OR TOPIC: (SARS-CoV-2)

Refined by: DOCUMENT TYPES: (ARTICLE)

Timespan: All years. Indexes: SCI-EXPANDED.

TOTAL: **40,765 Articles; 340,868 Authorships.**

Fields (authors, title, journal, year, etc) extracted with the BIBEXCEL software: https://homepage.univie.ac.at/juan.gorraiz/bibexcel/

Persson, O., R. Danell, J. Wiborg Schneider. 2009. How to use Bibexcel for various types of bibliometric analysis. In Celebrating scholarly communication studies: A Festschrift for Olle Persson at his 60th Birthday, ed. F. Åström, R. Danell, B. Larsen, J. Schneider, p 9–24. Leuven, Belgium: International Society for Scientometrics and Informetrics.

**Gender of Authorships** identified (92.2%) by means of the following databases:

GenderChecker: https://genderchecker.com/
GenderAPI: https://genderapi.io/
and Google Search / Images.

Table S1. All data organized by countries (according with the corresponding address).

| | Articles | % | Authorships | Authors/ Article | Known Gender | Men | % | Women | % |
|---|---|---|---|---|---|---|---|---|---|
| Afghanistan | 3 | 0.0 | 12 | 4.00 | 10 | 9 | 90.0 | 1 | 10.0 |
| Albania | 8 | 0.0 | 43 | 5.38 | 40 | 13 | 32.5 | 27 | 67.5 |
| Algeria | 24 | 0.1 | 147 | 6.13 | 121 | 65 | 53.7 | 56 | 46.3 |
| Andorra | 1 | 0.0 | 13 | 13.00 | 12 | 6 | 50.0 | 6 | 50.0 |
| Angola | 1 | 0.0 | 14 | 14.00 | 13 | 5 | 38.5 | 8 | 61.5 |
| Argentina | 100 | 0.2 | 862 | 8.62 | 732 | 410 | 56.0 | 322 | 44.0 |
| Australia | 836 | 2.1 | 6196 | 7.41 | 5946 | 3440 | 57.9 | 2506 | 42.1 |
| Austria | 221 | 0.5 | 1990 | 9.00 | 1866 | 1251 | 67.0 | 615 | 33.0 |
| Azerbaijan | 3 | 0.0 | 6 | 2.00 | 2 | 2 | 100.0 | 0 | 0.0 |
| Bahrain | 11 | 0.0 | 72 | 6.55 | 71 | 45 | 63.4 | 26 | 36.6 |
| Bangladesh | 134 | 0.3 | 782 | 5.84 | 701 | 542 | 77.3 | 159 | 22.7 |
| Barbados | 1 | 0.0 | 1 | 1.00 | 1 | 1 | 100.0 | 0 | 0.0 |
| BELARUS | 4 | 0.0 | 17 | 4.25 | 17 | 12 | 70.6 | 5 | 29.4 |
| Belgium | 299 | 0.7 | 2795 | 9.35 | 2539 | 1595 | 62.8 | 944 | 37.2 |
| Belize | 1 | 0.0 | 2 | 2.00 | 2 | 2 | 100.0 | 0 | 0.0 |
| Benin | 3 | 0.0 | 19 | 6.33 | 16 | 14 | 87.5 | 2 | 12.5 |
| Bhutan | 3 | 0.0 | 15 | 5.00 | 13 | 9 | 69.2 | 4 | 30.8 |
| Bolivia | 2 | 0.0 | 21 | 10.50 | 21 | 12 | 57.1 | 9 | 42.9 |
| Bosnia & Herceg | 12 | 0.0 | 72 | 6.00 | 70 | 32 | 45.7 | 38 | 54.3 |
| Botswana | 1 | 0.0 | 3 | 3.00 | 3 | 3 | 100.0 | 0 | 0.0 |
| Brazil | 920 | 2.3 | 7168 | 7.79 | 6928 | 3704 | 53.5 | 3224 | 46.5 |
| Brunei | 11 | 0.0 | 61 | 5.55 | 59 | 37 | 62.7 | 22 | 37.3 |
| Bulgaria | 14 | 0.0 | 51 | 3.64 | 49 | 25 | 51.0 | 24 | 49.0 |
| Burkina Faso | 2 | 0.0 | 16 | 8.00 | 3 | 2 | 66.7 | 1 | 33.3 |

| | | | | | | | | | |
|---|---|---|---|---|---|---|---|---|---|
| Burundi | 1 | 0.0 | 10 | 10.00 | 1 | 0 | 0.0 | 1 | 100.<br>0 |
| Cambodia | 2 | 0.0 | 10 | 5.00 | 8 | 7 | 87.5 | 1 | 12.5 |
| Cameroon | 8 | 0.0 | 64 | 8.00 | 46 | 22 | 47.8 | 24 | 52.2 |
| Canada | 972 | 2.4 | 7002 | 7.20 | 6688 | 3848 | 57.5 | 2840 | 42.5 |
| Chile | 121 | 0.3 | 857 | 7.08 | 821 | 508 | 61.9 | 313 | 38.1 |
| Colombia | 85 | 0.2 | 573 | 6.74 | 567 | 379 | 66.8 | 188 | 33.2 |
| Costa Rica | 4 | 0.0 | 19 | 4.75 | 19 | 6 | 31.6 | 13 | 68.4 |
| Croatia | 60 | 0.1 | 409 | 6.82 | 382 | 196 | 51.3 | 186 | 48.7 |
| Cuba | 14 | 0.0 | 181 | 12.93 | 150 | 67 | 44.7 | 83 | 55.3 |
| Cyprus | 31 | 0.1 | 178 | 5.74 | 174 | 115 | 66.1 | 59 | 33.9 |
| Czech Republic | 75 | 0.2 | 499 | 6.65 | 465 | 285 | 61.3 | 180 | 38.7 |
| Dem Rep Congo | 10 | 0.0 | 101 | 10.10 | 97 | 77 | 79.4 | 20 | 20.6 |
| Denmark | 161 | 0.4 | 1742 | 10.82 | 1696 | 1022 | 60.3 | 674 | 39.7 |
| Djibouti | 1 | 0.0 | 3 | 3.00 | 3 | 3 | 100.0 | 0 | 0.0 |
| Dominican Rep | 3 | 0.0 | 9 | 3.00 | 8 | 4 | 50.0 | 4 | 50.0 |
| Ecuador | 32 | 0.1 | 206 | 6.44 | 202 | 129 | 63.9 | 73 | 36.1 |
| Egypt | 275 | 0.7 | 1538 | 5.59 | 1463 | 870 | 59.5 | 593 | 40.5 |
| Estonia | 10 | 0.0 | 71 | 7.10 | 70 | 41 | 58.6 | 29 | 41.4 |
| Ethiopia | 112 | 0.3 | 755 | 6.74 | 523 | 420 | 80.3 | 103 | 19.7 |
| Fiji | 1 | 0.0 | 2 | 2.00 | 2 | 1 | 50.0 | 1 | 50.0 |
| Finland | 80 | 0.2 | 637 | 7.96 | 565 | 295 | 52.2 | 270 | 47.8 |
| France | 1189 | 2.9 | 13603 | 11.44 | 12192 | 7556 | 62.0 | 4636 | 38.0 |
| French Guiana | 5 | 0.0 | 63 | 12.60 | 63 | 29 | 46.0 | 34 | 54.0 |
| Gabon | 2 | 0.0 | 12 | 6.00 | 9 | 5 | 55.6 | 4 | 44.4 |
| Gambia | 1 | 0.0 | 3 | 3.00 | 3 | 0 | 0.0 | 3 | 100.<br>0 |
| Georgia | 2 | 0.0 | 9 | 4.50 | 9 | 7 | 77.8 | 2 | 22.2 |
| Germany | 1458 | 3.6 | 13818 | 9.48 | 12680 | 8675 | 68.4 | 4005 | 31.6 |
| Ghana | 26 | 0.1 | 196 | 7.54 | 194 | 154 | 79.4 | 40 | 20.6 |

| | | | | | | | | | |
|---|---|---|---|---|---|---|---|---|---|
| Gibraltar | 3 | 0.0 | 9 | 3.00 | 9 | 6 | 66.7 | 3 | 33.3 |
| Greece | 233 | 0.6 | 2211 | 9.49 | 2076 | 1262 | 60.8 | 814 | 39.2 |
| Guinea | 1 | 0.0 | 11 | 11.00 | 10 | 9 | 90.0 | 1 | 10.0 |
| Guyana | 1 | 0.0 | 7 | 7.00 | 7 | 5 | 71.4 | 2 | 28.6 |
| Haiti | 1 | 0.0 | 15 | 15.00 | 14 | 10 | 71.4 | 4 | 28.6 |
| Honduras | 2 | 0.0 | 9 | 4.50 | 5 | 4 | 80.0 | 1 | 20.0 |
| Hungary | 99 | 0.2 | 717 | 7.24 | 676 | 460 | 68.0 | 216 | 32.0 |
| Iceland | 9 | 0.0 | 93 | 10.33 | 88 | 54 | 61.4 | 34 | 38.6 |
| India | 1703 | 4.2 | 10104 | 5.93 | 8864 | 6277 | 70.8 | 2587 | 29.2 |
| Indonesia | 81 | 0.2 | 530 | 6.54 | 463 | 254 | 54.9 | 209 | 45.1 |
| Iran | 620 | 1.5 | 4249 | 6.85 | 4099 | 2558 | 62.4 | 1541 | 37.6 |
| Iraq | 27 | 0.1 | 99 | 3.67 | 90 | 73 | 81.1 | 17 | 18.9 |
| Ireland | 151 | 0.4 | 1096 | 7.26 | 1021 | 566 | 55.4 | 455 | 44.6 |
| Israel | 348 | 0.9 | 2318 | 6.66 | 2249 | 1367 | 60.8 | 882 | 39.2 |
| Italy | 3174 | 7.8 | 33297 | 10.49 | 31478 | 17978 | 57.1 | 13500 | 42.9 |
| Jamaica | 1 | 0.0 | 4 | 4.00 | 4 | 0 | 0.0 | 4 | 100.0 |
| Japan | 648 | 1.6 | 5217 | 8.05 | 5046 | 3935 | 78.0 | 1111 | 22.0 |
| Jordan | 84 | 0.2 | 458 | 5.45 | 452 | 268 | 59.3 | 184 | 40.7 |
| Kazakhstan | 10 | 0.0 | 62 | 6.20 | 57 | 40 | 70.2 | 17 | 29.8 |
| Kenya | 18 | 0.0 | 124 | 6.89 | 119 | 81 | 68.1 | 38 | 31.9 |
| Kosovo | 1 | 0.0 | 6 | 6.00 | 6 | 6 | 100.0 | 0 | 0.0 |
| Kuwait | 33 | 0.1 | 216 | 6.55 | 214 | 146 | 68.2 | 68 | 31.8 |
| Laos | 1 | 0.0 | 14 | 14.00 | 13 | 8 | 61.5 | 5 | 38.5 |
| Latvia | 7 | 0.0 | 39 | 5.57 | 39 | 21 | 53.8 | 18 | 46.2 |
| Lebanon | 56 | 0.1 | 374 | 6.68 | 365 | 202 | 55.3 | 163 | 44.7 |
| Libya | 13 | 0.0 | 187 | 14.38 | 175 | 102 | 58.3 | 73 | 41.7 |
| Liechtenstein | 3 | 0.0 | 26 | 8.67 | 22 | 18 | 81.8 | 4 | 18.2 |
| Lithuania | 16 | 0.0 | 84 | 5.25 | 83 | 33 | 39.8 | 50 | 60.2 |
| Luxembourg | 8 | 0.0 | 67 | 8.38 | 66 | 42 | 63.6 | 24 | 36.4 |

| | | | | | | | | | |
|---|---|---|---|---|---|---|---|---|---|
| Madagascar | 2 | 0.0 | 15 | 7.50 | 11 | 7 | 63.6 | 4 | 36.4 |
| Malawi | 4 | 0.0 | 38 | 9.50 | 36 | 29 | 80.6 | 7 | 19.4 |
| Malaysia | 156 | 0.4 | 1011 | 6.48 | 910 | 571 | 62.7 | 339 | 37.3 |
| Maldives | 2 | 0.0 | 8 | 4.00 | 5 | 2 | 40.0 | 3 | 60.0 |
| Mali | 1 | 0.0 | 8 | 8.00 | 1 | 1 | 100.0 | 0 | 0.0 |
| Malta | 17 | 0.0 | 72 | 4.24 | 67 | 39 | 58.2 | 28 | 41.8 |
| Mauritius | 6 | 0.0 | 20 | 3.33 | 19 | 13 | 68.4 | 6 | 31.6 |
| Mexico | 273 | 0.7 | 1967 | 7.21 | 1860 | 1234 | 66.3 | 626 | 33.7 |
| Moldova | 1 | 0.0 | 9 | 9.00 | 8 | 5 | 62.5 | 3 | 37.5 |
| Monaco | 1 | 0.0 | 14 | 14.00 | 14 | 11 | 78.6 | 3 | 21.4 |
| Montenegro | 3 | 0.0 | 15 | 5.00 | 15 | 8 | 53.3 | 7 | 46.7 |
| Morocco | 71 | 0.2 | 456 | 6.42 | 402 | 269 | 66.9 | 133 | 33.1 |
| Mozambique | 3 | 0.0 | 48 | 16.00 | 48 | 20 | 41.7 | 28 | 58.3 |
| Nepal | 62 | 0.2 | 291 | 4.69 | 274 | 193 | 70.4 | 81 | 29.6 |
| Netherlands | 363 | 0.9 | 3057 | 8.42 | 2740 | 1615 | 58.9 | 1125 | 41.1 |
| New Caledonia | 1 | 0.0 | 6 | 6.00 | 6 | 4 | 66.7 | 2 | 33.3 |
| New Zealand | 90 | 0.2 | 592 | 6.58 | 562 | 269 | 47.9 | 293 | 52.1 |
| Nicaragua | 1 | 0.0 | 6 | 6.00 | 6 | 4 | 66.7 | 2 | 33.3 |
| Nigeria | 101 | 0.2 | 631 | 6.25 | 510 | 359 | 70.4 | 151 | 29.6 |
| North Macedonia | 3 | 0.0 | 12 | 4.00 | 12 | 9 | 75.0 | 3 | 25.0 |
| Norway | 136 | 0.3 | 952 | 7.00 | 907 | 531 | 58.5 | 376 | 41.5 |
| Oman | 28 | 0.1 | 228 | 8.14 | 190 | 115 | 60.5 | 75 | 39.5 |
| Pakistan | 277 | 0.7 | 1425 | 5.14 | 1357 | 911 | 67.1 | 446 | 32.9 |
| Palestine | 16 | 0.0 | 68 | 4.25 | 67 | 33 | 49.3 | 34 | 50.7 |
| Panama | 3 | 0.0 | 22 | 7.33 | 21 | 18 | 85.7 | 3 | 14.3 |
| Paraguay | 4 | 0.0 | 27 | 6.75 | 27 | 18 | 66.7 | 9 | 33.3 |
| Peoples R China | 5861 | 14.4 | 54342 | 9.27 | 47654 | 28229 | 59.2 | 19425 | 40.8 |
| Peru | 38 | 0.1 | 221 | 5.82 | 216 | 154 | 71.3 | 62 | 28.7 |
| Philippines | 27 | 0.1 | 152 | 5.63 | 134 | 85 | 63.4 | 49 | 36.6 |
| Poland | 313 | 0.8 | 1740 | 5.56 | 1693 | 938 | 55.4 | 755 | 44.6 |

| | | | | | | | | | |
|---|---|---|---|---|---|---|---|---|---|
| Portugal | 180 | 0.4 | 1214 | 6.74 | 1140 | 536 | 47.0 | 604 | 53.0 |
| Qatar | 70 | 0.2 | 516 | 7.37 | 472 | 319 | 67.6 | 153 | 32.4 |
| Rep Congo | 5 | 0.0 | 47 | 9.40 | 46 | 27 | 58.7 | 19 | 41.3 |
| Romania | 151 | 0.4 | 893 | 5.91 | 871 | 414 | 47.5 | 457 | 52.5 |
| Russia | 148 | 0.4 | 1316 | 8.89 | 967 | 452 | 46.7 | 515 | 53.3 |
| Rwanda | 4 | 0.0 | 24 | 6.00 | 22 | 15 | 68.2 | 7 | 31.8 |
| Samoa | 1 | 0.0 | 3 | 3.00 | 3 | 1 | 33.3 | 2 | 66.7 |
| Saudi Arabia | 462 | 1.1 | 2794 | 6.05 | 2626 | 1926 | 73.3 | 700 | 26.7 |
| Senegal | 4 | 0.0 | 42 | 10.50 | 32 | 20 | 62.5 | 12 | 37.5 |
| Serbia | 66 | 0.2 | 445 | 6.74 | 432 | 199 | 46.1 | 233 | 53.9 |
| Sierra Leone | 2 | 0.0 | 10 | 5.00 | 10 | 8 | 80.0 | 2 | 20.0 |
| Singapore | 330 | 0.8 | 2933 | 8.89 | 2802 | 1729 | 61.7 | 1073 | 38.3 |
| Slovakia | 28 | 0.1 | 112 | 4.00 | 97 | 46 | 47.4 | 51 | 52.6 |
| Slovenia | 41 | 0.1 | 232 | 5.66 | 213 | 106 | 49.8 | 107 | 50.2 |
| Solomon Islands | 1 | 0.0 | 7 | 7.00 | 7 | 4 | 57.1 | 3 | 42.9 |
| Somalia | 2 | 0.0 | 13 | 6.50 | 13 | 10 | 76.9 | 3 | 23.1 |
| South Africa | 199 | 0.5 | 1223 | 6.15 | 795 | 509 | 64.0 | 286 | 36.0 |
| South Australia | 1 | 0.0 | 6 | 6.00 | 1 | 1 | 100.0 | 0 | 0.0 |
| South Korea | 659 | 1.6 | 4448 | 6.75 | 4208 | 2737 | 65.0 | 1471 | 35.0 |
| Spain | 1529 | 3.8 | 14091 | 9.22 | 13178 | 7051 | 53.5 | 6127 | 46.5 |
| Sri Lanka | 24 | 0.1 | 120 | 5.00 | 82 | 51 | 62.2 | 31 | 37.8 |
| Sudan | 10 | 0.0 | 67 | 6.70 | 63 | 40 | 63.5 | 23 | 36.5 |
| Sweden | 257 | 0.6 | 2080 | 8.09 | 1960 | 1174 | 59.9 | 786 | 40.1 |
| Switzerland | 399 | 1.0 | 3230 | 8.10 | 3058 | 1906 | 62.3 | 1152 | 37.7 |
| Syria | 5 | 0.0 | 14 | 2.80 | 13 | 9 | 69.2 | 4 | 30.8 |
| Taiwan | 316 | 0.8 | 2091 | 6.62 | 1985 | 1251 | 63.0 | 734 | 37.0 |
| Tanzania | 3 | 0.0 | 7 | 2.33 | 6 | 5 | 83.3 | 1 | 16.7 |
| Thailand | 115 | 0.3 | 867 | 7.54 | 772 | 430 | 55.7 | 342 | 44.3 |
| Togo | 2 | 0.0 | 39 | 19.50 | 35 | 25 | 71.4 | 10 | 28.6 |
| Trinidad Tobago | 2 | 0.0 | 10 | 5.00 | 9 | 6 | 66.7 | 3 | 33.3 |

| | | | | | | | | | |
|---|---|---|---|---|---|---|---|---|---|
| Tunisia | 34 | 0.1 | 399 | 11.74 | 398 | 222 | 55.8 | 176 | 44.2 |
| Turkey | 1011 | 2.5 | 5927 | 5.86 | 5744 | 3476 | 60.5 | 2268 | 39.5 |
| U Arab Emirates | 116 | 0.3 | 724 | 6.24 | 684 | 448 | 65.5 | 236 | 34.5 |
| Uganda | 16 | 0.0 | 123 | 7.69 | 116 | 86 | 74.1 | 30 | 25.9 |
| UK | 2372 | 5.8 | 24732 | 10.43 | 20298 | 11930 | 58.8 | 8368 | 41.2 |
| Ukraine | 8 | 0.0 | 54 | 6.75 | 37 | 24 | 64.9 | 13 | 35.1 |
| Uruguay | 9 | 0.0 | 65 | 7.22 | 52 | 28 | 53.8 | 24 | 46.2 |
| USA | 9274 | 22.8 | 75875 | 8.18 | 73267 | 42665 | 58.2 | 30602 | 41.8 |
| Uzbekistan | 1 | 0.0 | 5 | 5.00 | 4 | 1 | 25.0 | 3 | 75.0 |
| Vatican | 4 | 0.0 | 19 | 4.75 | 17 | 14 | 82.4 | 3 | 17.6 |
| Venezuela | 7 | 0.0 | 46 | 6.57 | 43 | 23 | 53.5 | 20 | 46.5 |
| Vietnam | 97 | 0.2 | 824 | 8.49 | 781 | 516 | 66.1 | 265 | 33.9 |
| Yemen | 12 | 0.0 | 71 | 5.92 | 69 | 55 | 79.7 | 14 | 20.3 |
| Zambia | 2 | 0.0 | 44 | 22.00 | 38 | 32 | 84.2 | 6 | 15.8 |
| Zimbabwe | 11 | 0.0 | 38 | 3.45 | 35 | 28 | 80.0 | 7 | 20.0 |
